# Inflammatory Biomarkers and Physiomarkers of Late-Onset Sepsis and Necrotizing Enterocolitis in Premature Infants

**DOI:** 10.1101/2023.06.29.23292047

**Authors:** Rupin Kumar, Sherry Kausch, Angela K.S. Gummadi, Karen D. Fairchild, Mayuresh Abhyankar, William A. Petri, Brynne A. Sullivan

## Abstract

**Impact:**

- Late-onset sepsis and necrotizing enterocolitis (NEC) in very low birth weight (VLBW, <1500g) premature infants can result in severe morbidity and mortality. Diagnosis is challenging due to overlap with non-infectious conditions, leading to a delayed or unnecessary antibiotic use.
- In a single-center cohort of VLBW infants, inflammatory biomarkers were elevated at the time of sepsis due to Gram-negative sepsis or NEC, but not other sepsis; compared to times without sepsis or NEC.
- Physiomarkers of sepsis correlate with some biomarkers of sepsis, and combining their information could help in the early diagnosis of sepsis.

**Background:** Early diagnosis of late-onset sepsis (LOS) and necrotizing enterocolitis (NEC) in VLBW (<1500g) infants is challenging due to non-specific clinical signs. Inflammatory biomarkers increase in response to infection, but non-infectious conditions also cause inflammation in premature infants. Physiomarkers of sepsis exist in cardiorespiratory data and may be useful in combination with biomarkers for early diagnosis.

**Objectives:** To determine whether inflammatory biomarkers at LOS or NEC diagnosis differ from times without infection, and whether biomarkers correlate with a cardiorespiratory physiomarker score.

**Methods:** We collected remnant plasma samples and clinical data from VLBW infants. Sample collection occurred with blood draws for routine laboratory testing and blood draws for suspected sepsis. We analyzed 11 inflammatory biomarkers and a continuous cardiorespiratory monitoring (POWS) score. We compared biomarkers at gram-negative (GN) bacteremia or NEC, gram-positive (GP) bacteremia, negative blood cultures, and routine samples.

**Results:** We analyzed 188 samples in 54 VLBW infants. Biomarker levels varied widely, even at routine laboratory testing. Several biomarkers were increased at the time of GN LOS or NEC diagnosis compared with all other samples. POWS was higher in patients with LOS and correlated with five biomarkers. IL-6 had 78% specificity at 100% sensitivity to detect GN LOS or NEC and added information to POWS (AUC POWS = 0.610, POWS + IL-6 = 0.680).

**Conclusion(s):** Inflammatory biomarkers discriminate sepsis due to GN bacteremia or NEC and correlate with cardiorespiratory physiomarkers. Baseline biomarkers did not differ from times of GP bacteremia diagnosis or negative blood cultures.

## Introduction

Sepsis and necrotizing enterocolitis (NEC) lead to significant morbidity and mortality in premature infants^1^. Inflammation due to sepsis and NEC can cause end-organ damage^2, 3^, including brain injury, which leads to long-term impairment.^4–6^ Early recognition and treatment may improve outcomes, but signs and symptoms often overlap with non-infectious conditions^7^. Therefore, a diagnosis is often made once the infection is advanced, and on the other hand, empiric antibiotics are given when there is no infection.^8^ Clinicians have limited tests and data to guide decisions on the likelihood of infection.

Inflammatory molecules, upregulated as part of the immune response to infection, hold promise as biomarkers of neonatal sepsis. A biomarker with sufficient diagnostic accuracy could be a useful screening tool to aid in deciding whether to start antibiotics or continue monitoring for further signs of sepsis. Many studies have evaluated biomarkers for neonatal sepsis in premature infants.^9–12^ But, few have been translated into clinical care.^13^ Decisions to start or stop antibiotics in the NICU largely rely on the clinician’s assessment of clinical signs and non-specific laboratory tests, such as C-reactive protein^14^ and complete blood count (CBC) components.^15^

Physiologic markers of sepsis can be detected using predictive analysis of continuous vital sign data from standard bedside monitors.^16^ We used predictive analytics and a multicenter cohort of very low birth weight (VLBW, birth weight < 1500g) preterm infants to develop and externally validate a cardiorespiratory sepsis risk model. The model, called Pulse Oximetry Warning Score (POWS), detects abnormal heart rate and SpO2 patterns that occur near the clinical diagnosis of late-onset sepsis and calculates the relative risk of positive blood culture with sepsis in the subsequent 24 hours.^17^ We hypothesize that inflammatory biomarkers will correlate with physiologic markers of sepsis. Combining information from sepsis biomarkers and physiomarkers could add information on the likelihood of sepsis when vital sign-based predictive analytics detect a rising risk of sepsis.

While many prior studies have analyzed inflammatory biomarkers in premature infants at the time of suspected sepsis,^10, 18–20^ few have analyzed baseline levels or changes over time in patients who develop sepsis.^21, 22^ Inflammatory biomarkers may be less useful in premature infants with non-infectious conditions that lead to inflammation, including respiratory distress syndrome and chronic lung disease. To understand the potential utility of biomarkers in this unique population, more data are needed. In this study, we aim to identify inflammatory biomarkers that discriminate sepsis from baseline levels and sepsis-like illnesses in VLBW infants. To do this, we measured multiple biomarkers in samples collected from remnant plasma, weekly when available and at the time of evaluation for late-onset sepsis or NEC.

## Methods

### Study Design

This was a prospective, observational cohort study conducted at a single center Level IV academic NICU. The Institutional Review Board approved this study with verbal informed consent and a signed HIPAA data use authorization from a parent. We prospectively enrolled VLBW infants within seven days of birth and collected clinical data and remnant plasma from blood drawn for suspected sepsis and for routine laboratory testing. Clinical data and culture results were recorded from the electronic health record. We classified samples according to the indication for the laboratory tests ordered and the final diagnosis of a workup for LOS or NEC (see “clinical definitions” section below).

### Sample collection

The study protocol did not require any extra blood draws and did not influence the clinical team’s decisions on ordering blood tests. Nurses collected blood samples in tubes according to the clinical laboratory guidelines for the laboratory test ordered. Blood for a complete blood count was collected in EDTA tubes, and blood for a basic metabolic panel or C-reactive protein was collected in sodium heparin tubes. These were the only clinical laboratory tests used to request remnant plasma for the study. The clinical lab stores samples at 4 degrees Celsius immediately after receipt, and the leftover blood from a clinical sample was requested by the study team. The clinical lab separated plasma from whole blood for samples collected in sodium heparin tubes, while the study team separated plasma from whole blood for samples collected in EDTA tubes by centrifugation (1800 g X 5 min). We stored all remnant plasma samples with at least 250 microliters at −80C until analysis.

### Biomarker analysis

We selected subjects with at least two collected samples of adequate plasma volume (≥ 250 µL) for biomarker analysis. The multiplex assay measured a panel of inflammatory biomarkers: interleukin (IL)-6, IL-8, IL-10, IL-18, interferon gamma inducible protein (IP)-10, tumor necrosis factor alpha (TNFα), procalcitonin (PCT), human growth factor (HGF), endothelin growth factor (EGF), soluble suppression of tumorigenicity 2 (sST-2), and IL-1 receptor antigen (IL1-ra). We selected these biomarkers based on their role in the inflammatory response to bacterial infections and the existing literature. Analyte concentrations were quantified using a customized multiplex Luminex® magnetic bead-based antibody assay (R&D Systems, Minneapolis, USA). Fluorescence signals for each biomarker bead region were analyzed on a Luminex®200, a dual-laser flow-based detection instrument. Concentrations below the lowest standard were recorded as the value of the lower limit of detection for statistical analyses. Based on preliminary data, we used a 10-fold dilution to analyze PCT, sST-2, and IL-1ra, and the remaining biomarkers did not require dilution. Results of C-reactive protein (CRP) measurement obtained for clinical use were obtained from the electronic health record.

### Clinical definitions

Samples were classified as “routine” if they were obtained from blood drawn for standard laboratory monitoring and not during a period of suspected infection. We classified samples obtained at the time of a blood culture using the diagnosis of the event according to the following definitions:

A. Late-onset sepsis (LOS): a positive blood culture obtained after 72 hours of age and treated with at least five days of intravenous antibiotics. These were further categorized as sepsis due to Gram-positive (GP) or Gram-negative (GN) bacteremia according to the organism identified by blood culture.
B. Necrotizing enterocolitis (NEC): radiographic evidence of necrotizing enterocolitis and clinical illness with or without a positive blood culture.
C. Clinical sepsis (CS): negative blood and urine cultures treated with at least five days of antibiotics for presumed infection due to clinical illness
D. Sepsis Ruled Out (SRO): a negative blood culture treated with fewer than five days of antibiotics.

### Cardiorespiratory sepsis risk prediction

We previously developed a multivariable model to predict sepsis using continuous heart rate (HR) and oxygen saturation (SpO_2_) data called the Pulse Oximetry Warning Score, or POWS.^17^ The POWS model calculates the mean, standard deviation, skewness, kurtosis, and cross-correlation of HR and SpO_2_ every 10 minutes and uses logistic regression at each window to predict the relative risk of LOS with a positive blood culture in the next 24 hours. We calculated hourly POWS values from continuous bedside monitoring data during the 12 hours preceding blood cultures. POWS scores were calculated after discharge and thus did not influence decisions about clinical care.

### Statistical analyses

We compared distributions of inflammatory biomarkers and POWS data using Kruskal-Wallis analysis across the four diagnosis groups, followed by a pairwise Wilcoxon test with corrections for multiple testing. The relationships between POWS and individual biomarker levels were assessed using univariate linear regression. The statistical significance of each model’s coefficient was adjusted for repeated measures using the Huber-White method. A p-value < 0.05 was considered statistically significant. The predictive performance of biomarkers and POWS were assessed individually or in combination using sensitivity, specificity at specific thresholds, and area under the receiver operator characteristics curve (AUC) using logistic regression to predict LOS or NEC. All statistical analysis was performed using RStudio (R Version 4.2.3, Vienna, Austria).

## Results

### Patients and samples

We enrolled 118 VLBW infants with parental consent. Of those enrolled, we had sufficient samples from 54 infants for analysis. The final cohort had a mean gestational age of 25.9 ± 1.9 weeks and a mean birth weight of 803 ± 222 grams. Clinical characteristics overall and comparing those with and without LOS or NEC are shown in Table 1. In total, 188 plasma samples were analyzed, including 67 obtained near the time of blood cultures for suspected LOS or NEC and 120 at the time of routine blood sampling. Seven biomarkers (IP-10, IL-6, IL-10, IL-18, TNFa, IL-8, PCT) were analyzed at all times, while four (HGF, EGF, sST-2, IL-1ra) were successfully measured 80% of the time (152 samples in 45 patients).

**Table 1.**
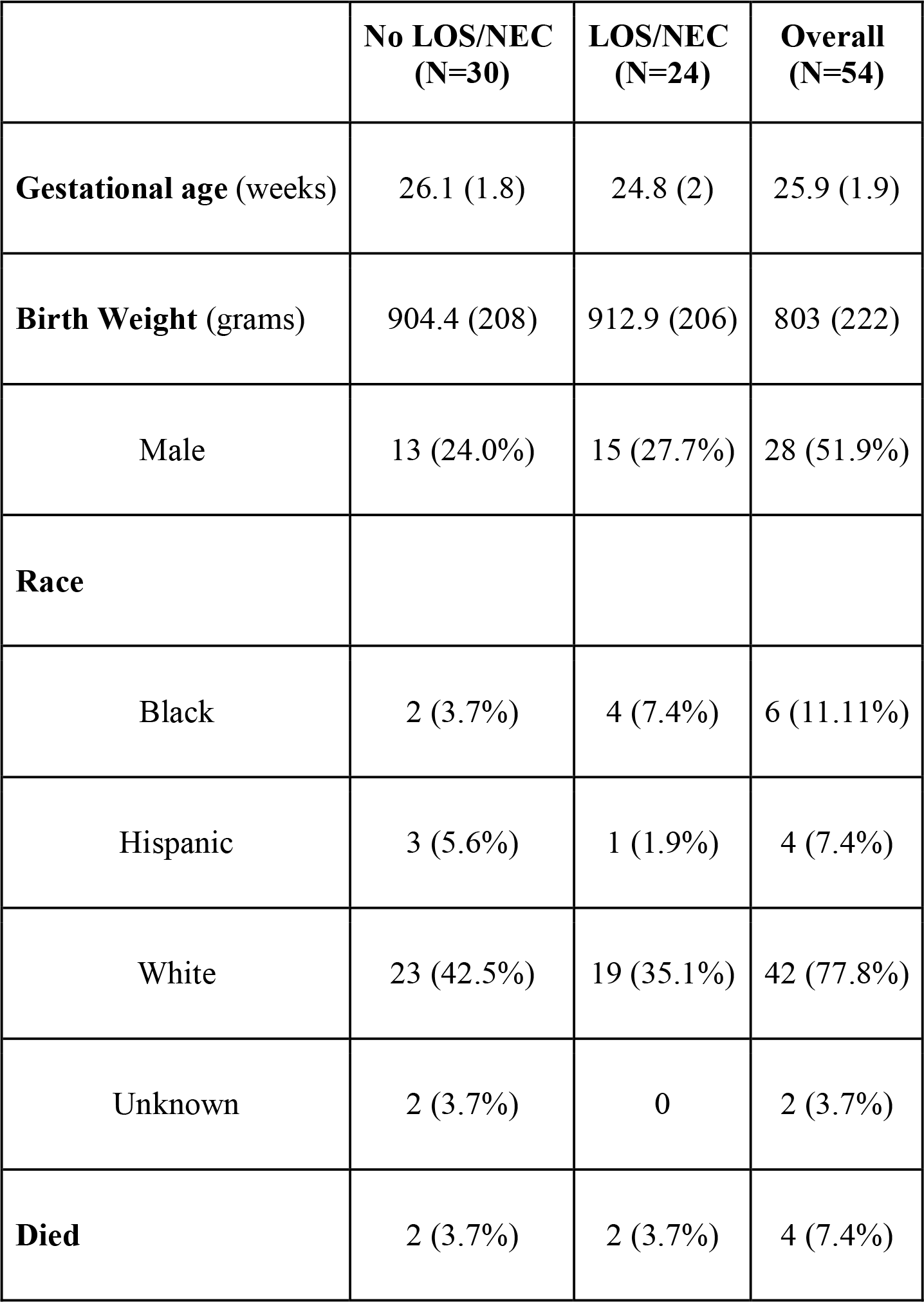
Cohort characteristics overall and grouped by infants with or with late-onset sepsis (LOS) or necrotizing enterocolitis (NEC) diagnosis. Values are presented as mean (standard deviation) or number (%).

|  | **No LOS/NEC**<br>**(N=30)** | **LOS/NEC**<br>**(N=24)** | **Overall**<br>**(N=54)** |
| --- | --- | --- | --- |
| **Gestational age** (weeks) | 26.1 (1.8) | 24.8 (2) | 25.9 (1.9) |
| **Birth Weight** (grams) | 904.4 (208) | 912.9 (206) | 803 (222) |
| Male | 13 (24.0%) | 15 (27.7%) | 28 (51.9%) |
| **Race** |  |  |  |
| Black | 2 (3.7%) | 4 (7.4%) | 6 (11.11%) |
| Hispanic | 3 (5.6%) | 1 (1.9%) | 4 (7.4%) |
| White | 23 (42.5%) | 19 (35.1%) | 42 (77.8%) |
| Unknown | 2 (3.7%) | 0 | 2 (3.7%) |
| **Died** | 2 (3.7%) | 2 (3.7%) | 4 (7.4%) |

Of the 67 blood cultures with study samples analyzed, 28 were diagnosed as LOS or NEC with bacteremia. Of these, there were 22 cases of Gram-positive (GP) bacteremia, 5 Gram-negative (GN) bacteremia, and 1 with NEC and GN bacteremia. Organisms in the positive blood cultures were coagulase-negative Staphylococcus (CONS) species (n=19), *Escherichia coli (n=2)*, *Enterobacter* species *(n=1)*, Group B *Streptococcus (n=1)*, *Klebsiella* species *(n=5)*, and methicillin-susceptible *Staphylococcus aureus (n=2)*. There were 2 cases of NEC with negative blood cultures and 17 negative blood cultures diagnosed as clinical sepsis (CS). The remaining 141 samples came from blood drawn at times with no sepsis (NS), including routine samples and blood cultures diagnosed as sepsis ruled out (SRO).

### Biomarkers

Five of the eleven biomarkers (IL-6, TNF-α, IL-8, IL-10, and sST-2) were significantly higher in patients with GN sepsis or NEC than those with NS, CS, and GP sepsis. Overall, biomarkers at CS or GP diagnosis were not significantly different from NS samples (**Figure 1**). In a few cases, infants with serial biomarker measurements and LOS or NEC had a rise from baseline at the time of diagnosis.

**Figure 1.**
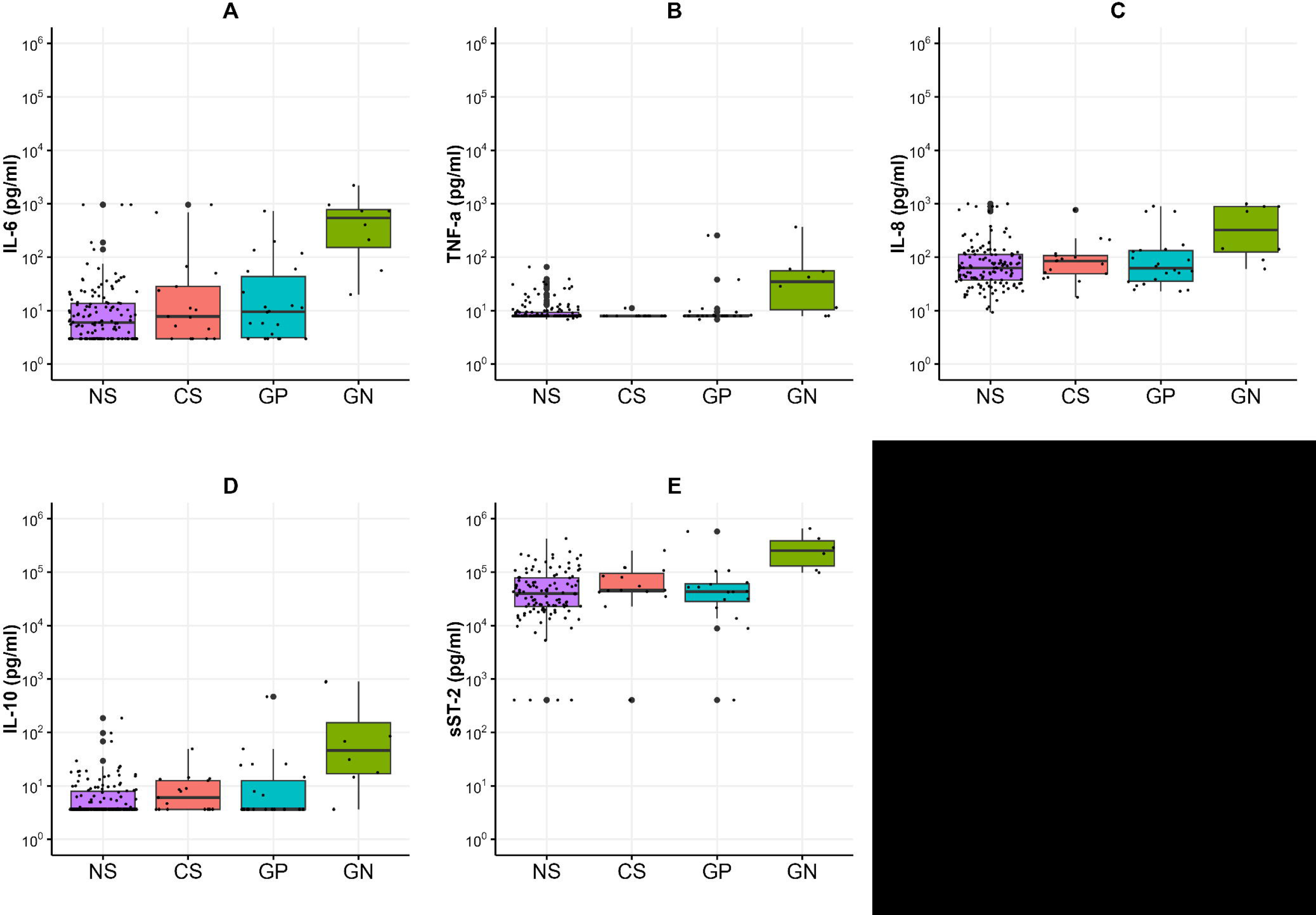
Plasma biomarker levels from remnant plasma collected at times with or without sepsis in VLBW infants. Samples were classified as clinical sepsis (CS, *n* = 17), Gram-positive bacteremia (GP, *n* = 22), Gram-negative bacteremia or NEC (GN, *n* = 8), or not sepsis (NS, *n* = 141). IL-6 (A), TNF-a (B), IL-8 (C), IL-10 (D), and sST-2 (E) were significantly higher in patients with GN sepsis as compared with NS, CS, and GP sepsis. Biomarker levels were not different when comparing NS, CS, and GP sepsis samples.

CRP was measured at the time of 92 samples (49%) and was highly variable. CRP was significantly higher in patients with GN sepsis or NEC (median CRP 4.4 g/dL, IQR 2.1 - 11.0) compared to patients with no sepsis (median CRP 0.1 g/dL, IQR 0.1 - 0.4) or clinical sepsis (median CRP 0.12 g/dL, IQR 0.1 - 1.2). There was no statistically significant difference in CRP for GN sepsis versus GP sepsis cases (median CRP 2.7 g/dL, IQR 0.1 - 3.3) or clinical sepsis (median CRP 0.12 g/dL, IQR 0.1 - 1.2).

Hierarchical cluster analysis of cytokines from 30 LOS or NEC blood samples showed two distinct clusters of biomarker profiles (**Figure 2**). GN bacteremia or NEC was more prevalent in the cluster of samples with the highest cytokine levels. IL-6 levels had the best overall test accuracy for diagnosing gram-negative sepsis or NEC with a specificity of 78% and a negative predictive value of 100% at a threshold of 200 pg/mL with 100% sensitivity.

**Figure 2.**
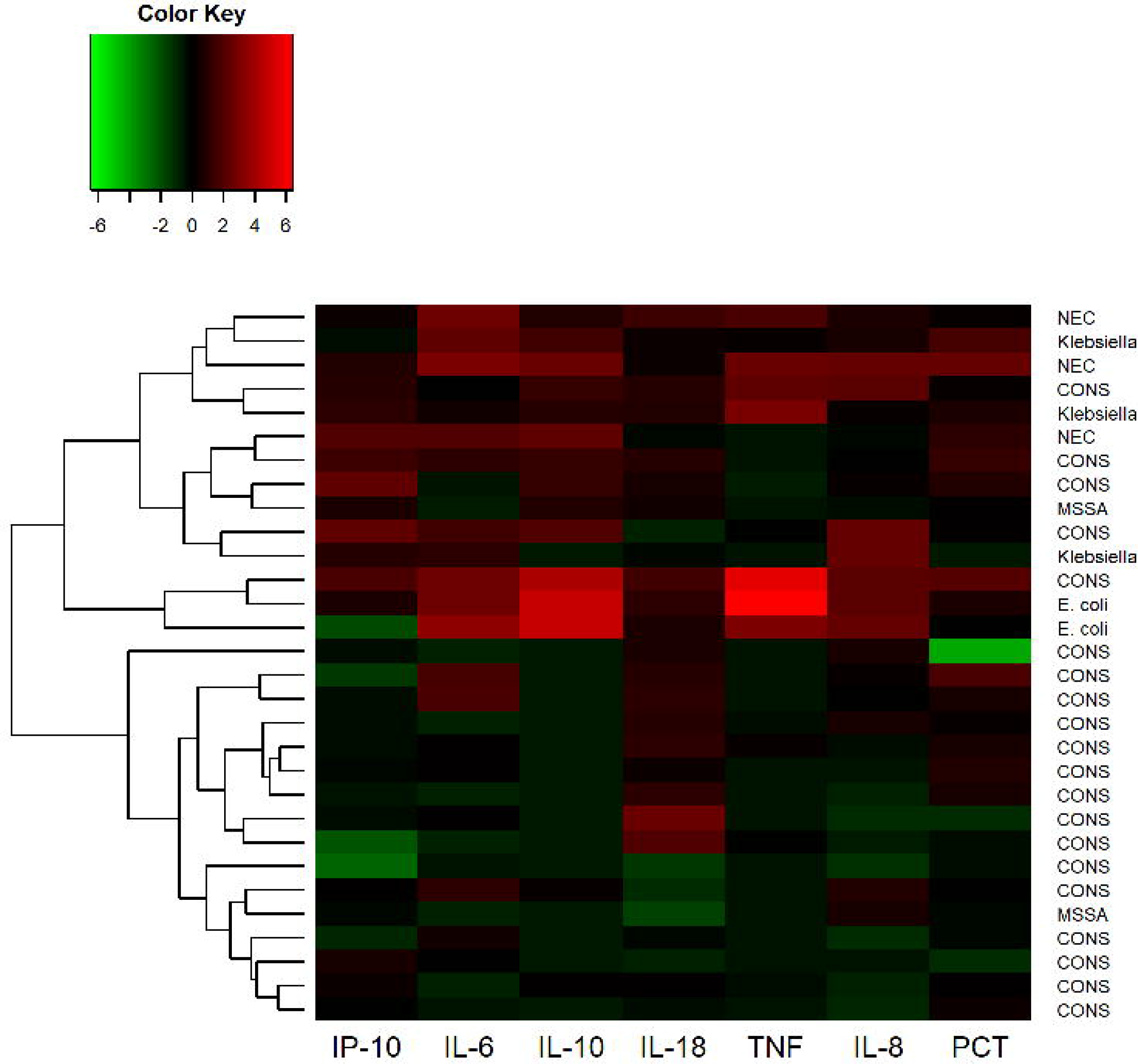
Hierarchical cluster analysis of cytokine levels in 30 cases of blood culture-positive sepsis and NEC. The scaled natural log of each cytokine was taken prior to clustering. Higher than average cytokine levels are depicted in shades of red and lower than average levels are depicted in green. The resulting hierarchical clustering dendrogram is on the left-hand side of the heatmap.

### Association with POWS, a cardiorespiratory sepsis risk prediction model

Of the 188 samples, 187 had continuous heart rate and oxygen saturation data available around the time of sample collection to calculate POWS, a pulse oximetry warning score designed as a physiomarker for impending sepsis or sepsis-like illness. POWS was significantly associated with the levels of 5 biomarkers (IL-8, PCT, HGF, sST-2, and IL1-ra) irrespective of the associated diagnosis (**Figure 3**, p<0.05). The maximum POWS within the 12 hours preceding the sample had an AUC to predict LOS (GN or GP) or NEC (with or without bacteremia) of 0.610. The AUC increased to 0.680 when the IL-6 level of the sample was added to the model.

**Figure 3.**
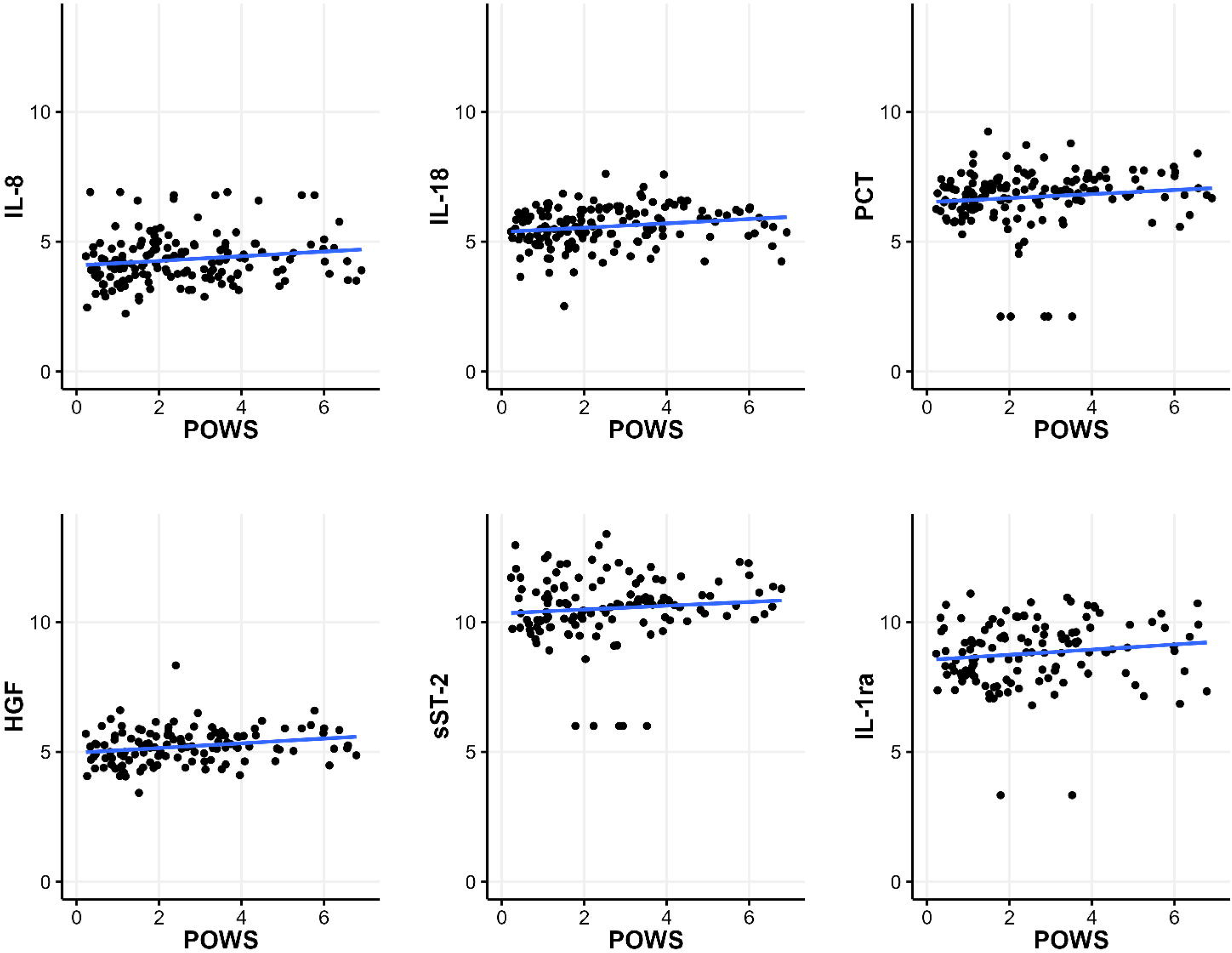
Relationship between POWS and inflammatory biomarkers. Plots of the scaled natural log of each biomarker level on the y-axis and the maximum sepsis risk within 12 hours before blood culture on the x-axis.

## Discussion

We assayed inflammatory biomarkers at the time of blood culture for suspected sepsis and routine laboratory testing in VLBW infants and found differences that distinguish late-onset Gram-negative sepsis and NEC from other diagnoses, but similar biomarker distributions in samples collected at baseline and at the time of suspected sepsis or Gram-positive sepsis. We also found correlations between inflammatory biomarkers and POWS, an algorithm that detects abnormal patterns of heart rate and oxygen saturation that we previously developed as a physiomarker of sepsis.

Our results confirm the findings in previous work that multiple biomarkers can discriminate between GN sepsis and GP sepsis at the time of positive blood culture.^12^ Since Gram-negative infections and NEC are known to carry higher morbidity and mortality, this finding might be a useful adjunct clinical decision support tool in helping pick the appropriate empiric antibiotic therapy and institute treatment early.^23–25^ The current study adds to our prior work because we analyzed samples remote from suspected sepsis. We found no significant difference in biomarker levels in these samples compared to those obtained at the time of clinical sepsis or Gram-positive sepsis compared to times when sepsis was not suspected or was ruled out. Some VLBW preterm infants may have a chronic or subacute systemic inflammatory response associated with lung disease or intracranial hemorrhage, which could confound sepsis prediction using blood biomarkers or cardiorespiratory physiomarkers.^26–28^

Plasma cytokines and chemokines are early markers of immune activation in response to infection, and have been shown to rise in septic premature infants.^22, 29, 30^ Several studies have evaluated the utility of biomarkers for diagnosing sepsis by comparing biomarker levels in cases versus controls^11^. Others have compared cytokine profiles obtained at the time of suspected infection in VLBW infants when the blood culture returns positive versus negative.^31, 32^ Instead, we assessed longitudinal changes in inflammatory biomarkers measured at times when there was clinical suspicion for infection or sepsis and at times remote from infection. Kuster and colleagues also took the approach of measuring cytokines at baseline and at the time of suspected sepsis in VLBW infants and found IL-1ra and IL-6 to have high sensitivity and specificity in 21 cases of late-onset sepsis, even measured from samples taken the day prior to blood culture.^30^ A longitudinal study of extremely low birth weight (<1000g) measured cytokines at days 1, 3, 14, and 21 after birth and found that IFN-γ, IL-10, IL-18, TGF-β, and TNF-α levels differed among infants who developed fungal or bacterial LOS compared with those who never developed sepsis.^22^ This study enrolled a large cohort from the NICHD Neonatal Research Network centers but did not evaluate differences relative to the timing of blood cultures. They found that infants with higher levels of immune regulatory cytokines relative to pro-inflammatory cytokines were associated with an increased risk of LOS at any time during the NICU course.^21^

Of the biomarkers assayed in our study, IL-6 has been most consistently identified as a promising sepsis biomarker in previous studies, though with variable sensitivity and specificity.^32^ We found that IL-6 had a high specificity at a threshold set to detect 100% of Gram-negative sepsis and NEC cases, but low predictive accuracy for Gram-positive cases, which were mostly due to CONS bacteremia. We also found highly elevated levels of IL-6 measured from samples remote from sepsis with blood draws for routine laboratory tests.

The inclusion of sST-2 as a sepsis biomarker was novel for this population as its utility has mainly been studied in adult populations. It is an IL-1 family receptor that binds IL-33 and has been implicated in diseases involving intestinal inflammation.^33^ Recent studies in adult patients demonstrate an association between elevated sST-2 and the severity of illness in *Clostridium difficile* colitis.^34, 35^ Our results indicate that this may also be a useful biomarker for diagnosing NEC and Gram-negative sepsis, where gastrointestinal dysfunction and bacterial translocation cause systemic inflammation.

Cytokines and chemokines such as IL-6, IL-1ra, and IL-8 have been demonstrated to have diagnostic utility as early sepsis markers ^30, 36^, while acute phase reactants such as CRP and PCT rise during the later phases of systemic inflammation.^37^ CRP and PCT are also examples of the few inflammatory biomarkers available in U.S. clinical laboratories, which may drive their clinical use despite evidence of low clinical utility.^14^ A recent meta-analysis showed overall low sensitivity and specificity of CRP for late-onset sepsis diagnosis.^38^

We note several limitations of the study. First, the analysis was limited by the small sample size, where most late-onset sepsis events were due to CONS bacteremia, some of which may have represented contamination and not a true infection. Second, events diagnosed as clinical sepsis are heterogeneous in severity with subjective diagnostic criteria. We collected data on clinical characteristics, but not to the level of detail to account for concurrent inflammatory processes, such as lung disease, invasive mechanical ventilation, and minor procedures. Finally, the use of remnant plasma allowed us to enroll patients and collect samples without additional blood draws, but the volume of plasma available was small and therefore did not allow assays to be run in duplicate.

Correlations with physiomarkers of sepsis using POWS, a cardiorespiratory sepsis risk score, resulted in promising correlations that warrant further confirmation in larger studies. Continuous cardiorespiratory predictive monitoring used in conjunction with biomarker testing could prove useful, both for early initiation of antibiotics when the likelihood of sepsis is high and for sparing antibiotics when biomarkers and physiomarkers indicate low sepsis risk.

## Conclusion

In conclusion, in a prospective cohort of VLBW infants, inflammatory biomarkers discriminated between late-onset sepsis due to gram-negative bacteremia or NEC and all other samples collected at baseline or times of suspected sepsis with negative blood culture or gram-positive bacteremia. Several inflammatory biomarkers, measured at baseline and at the time of suspected or confirmed sepsis, correlated with cardiorespiratory physiomarkers of sepsis. Baseline inflammatory biomarker levels did not differ from levels obtained at the time of sepsis due to gram-positive bacteremia or negative blood cultures.

## Data Availability

All data produced in the present study are available upon reasonable request to the authors.

## Acknowledgments

None

## Funding

We acknowledge the following grant for funding the work presented in this manuscript: K23 HD097254 (PI: B Sullivan); R01 AI124214, R01 AI152477 (PI: W. Petri)

## Author Contributions

All authors made substantial contributions to the conception, performance, or writing of the work; RK, SK, and BS made substantial contributions to the acquisition, analysis, or interpretation of data; RK, BS, and AG drafted the work and all other authors have substantively revised it. All authors have approved the submitted version. All authors have agreed both to be personally accountable for the author’s own contributions and to ensure that questions related to the accuracy or integrity of any part of the work, even ones in which the author was not personally involved, are appropriately investigated, resolved, and the resolution documented in the literature.

## Competing interests

The authors have no competing interests to disclose.

## Consent Statement

The Institutional Review Board of University of Virginia gave approval for this study with verbal informed consent and a signed HIPAA data use authorization from a parent.

## Notes

### Competing Interest Statement

The authors have declared no competing interest.

